# Performance of an Optimized Methylation-Protein Multi-Cancer Early Detection (MCED) Test Classifier

**DOI:** 10.64898/2026.03.03.26347329

**Authors:** Vladimir G. Gainullin, Melissa Gray, Madhav Kumar, Stephen Luebker, Amy M. Lehman, Omair A. Choudhry, Julia Roberta, Darl D. Flake, Avinash Shanmugam, Kevin Cortes, Emily Chang, Philip J. Uren, Amin Mazloom, Jorge Garces, Gerard A. Silvestri, David W. Chesla, Robert W. Given, Tomasz M. Beer, Frank Diehl

**Affiliations:** Exact Sciences Corporation, Madison, WI, USA; Exact Sciences Corporation, La Jolla, CA, USA; Medical University of South Carolina, Charleston, SC, USA; Corewell Health, Grand Rapids, MI, USA; Urology of Virginia, Virginia Beach, VA, USA

**Keywords:** Multi-cancer early detection, circulating tumor DNA (ctDNA), methylation, proteins, machine learning

## Abstract

Multi-cancer early detection (MCED) tests can detect several cancer types and stages. We previously developed a methylation and protein (MP V1) MCED classifier. In this study, we present a refined MP V2 classifier, developed by evaluating model architectures that improved performance in prospectively enrolled case-control cohorts under standard testing conditions. The newly developed MP V2 classifier was trained to be more generalizable and achieve increased early-stage sensitivity at a target specificity of ≥97.0%. MP V1 and MP V2 classifier performances were compared using a previously described test set, and MP V2 performance was also evaluated in a new independent clinical validation set. Compared to MP V1, the MP V2 classifier demonstrated a 7.3% increase in overall sensitivity, with sensitivity increases of 7.6%, 9.2%, and 8.3% for stages I, II, and stages I/II, respectively, in the intended use (breast and prostate cancers excluded) test set. In an independent validation intended use set, the MP V2 classifier showed an overall sensitivity of 55.6%, with sensitivities of 26.8%, 42.9%, and 34.8% for stages I, II, and stages I/II, respectively. In a case-control setting, the MP V2 classifier offered improved sensitivity for early-stage cancers at a lower specificity target.

## Introduction

Cancer is the second leading cause of death in the United States, with more than 2 million new cancer cases and over 626,000 deaths projected for 2026.^1^ Guideline-recommended single-cancer screening is available for only four cancers: breast, cervical, colorectal, and lung.^2–5^ Such screening has resulted in early cancer detection and contributed to improved clinical outcomes and mortality for these cancer types.^1^ However, guideline-recommended single-cancer screening addresses only about 1/3 of incident cancers and cancer related deaths,^6^ with 2/3 lacking recommended screening options.^1^ Finally, current single-cancer screening tests sacrifice specificity to optimize sensitivity, resulting in substantial false positive (FP) rates. The reported FP rates for guideline-recommended single-cancer screening tests range from 6.0% for colorectal (age-adjusted mt-sDNA) to 14.5% for cervical (Papanicolaou test) for each round of screening.^7–10^ Since the probability of obtaining a FP result increases with each round of recommended breast, cervical, colorectal, and lung cancer screening, the cumulative estimated lifetime FP risk associated with guideline-recommended single-cancer screening is considerable.^11, 12^ To address some of these limitations and improve upon current population-based single-cancer screening efforts, significant efforts are being made in the development of early-detection screening strategies that simultaneously detect multiple cancers in a single blood test.

Advances in sensitive high-throughput technologies and machine learning have led to the development of minimally invasive, blood-based, single and multi-biomarker class multicancer early detection (MCED) tests. Several MCED tests are under development to identify multiple cancers earlier by detecting shared cancer-derived DNA (methylation, fragmentation, mutations), proteins, or other biomarkers in the blood.^13–19^ Since MCED tests interrogate cancer signals originating from multiple organ systems they represent a potential paradigm shift in cancer screening, and several assay attributes must be considered to optimize test development.

First, MCED tests must expand the range of screen detected cancers. Second, MCED tests must demonstrate sensitivity to detect early-stage cancers, particularly cancers where early-stage diagnoses enable intervention with curative intent. Third, sensitivity must be balanced with a relatively high specificity. For MCED tests, this balance aims to maximize potential benefits (i.e., early-stage detection) while keeping harms from FP results acceptable. These harms may be further reduced by an efficient diagnostic strategy following a positive MCED test that minimizes the diagnostic burden for patients.

Sensitivity for early-stage solid tumors, when treatment with curative intent is available, is likely to have an outsized impact on the benefits of screening with MCED. Notably, in the <u>D</u>etecting cancers <u>E</u>arlier <u>T</u>hrough <u>E</u>lective mutation-based blood <u>C</u>ollection and <u>T</u>esting (DETECT-A) trial, the first prospective, interventional study to investigate MCED testing, all eight patients whose cancers were treated following detection via MCED testing and subsequent diagnosis at Stage I or II remain alive and cancer-free after >4 years of follow up.^20^ While of modest size, this observation highlights the feasibility of early detection of solid tumors using a multi-biomarker class MCED test, and provides early evidence that successful treatment can be reliably delivered. These observations highlight the importance of an emphasis on early-stage sensitivity in MCED development. ^19^

We previously developed a methylation and protein (MP V1) MCED classifier utilizing a prospectively collected case-control study of individuals aged 50-84, allowing the detection of a broad range of cancer organ types at all stages.^15^ The specificity target for MP V1 was set at a minimum of ≥98.0%. Subsequent validation using an independent test set confirmed that the MP V1 classifier achieved the specificity requirement, with observed sensitivities of 15.4%, 38.0%, and 26.1% for stage I, stage II, and stage I and II combined, respectively.^15^

Molecular diagnostic tests, including MCED tests, are often developed using case-control studies.^21, 22^ MCED tests are designed with high specificity targets to limit FP results, and the performance observed using case-control samples may not accurately represent real-world performance in a clinical setting.^17, 18, 21, 22^ To demonstrate clinical utility, MCED tests must be designed to detect cancers at early stages when they may be more amenable to curative intent. In this algorithm development study, we developed an optimized MP classifier architecture (MP V2) using a lower target specificity (≥97.0%) in a case-control setting to optimize sensitivity for early-stage cancers. We then validated the classifier performance using an independent testing set that more closely resembled the intended-use population.

## Results

### Classifier Selection using an Independent Mini-holdout (MHO) Test Set

MP V1 and MP V2 performance were first examined in MHO classifier selection test set samples from a previously untested cohort of Ascertaining Serial Cancer patients to Enable New Diagnostic 2 (ASCEND-2) study subjects (Figure 1). A series of comparisons were performed to examine changes in target specificity, classifier model architecture, and both together.

**Figure 1.**
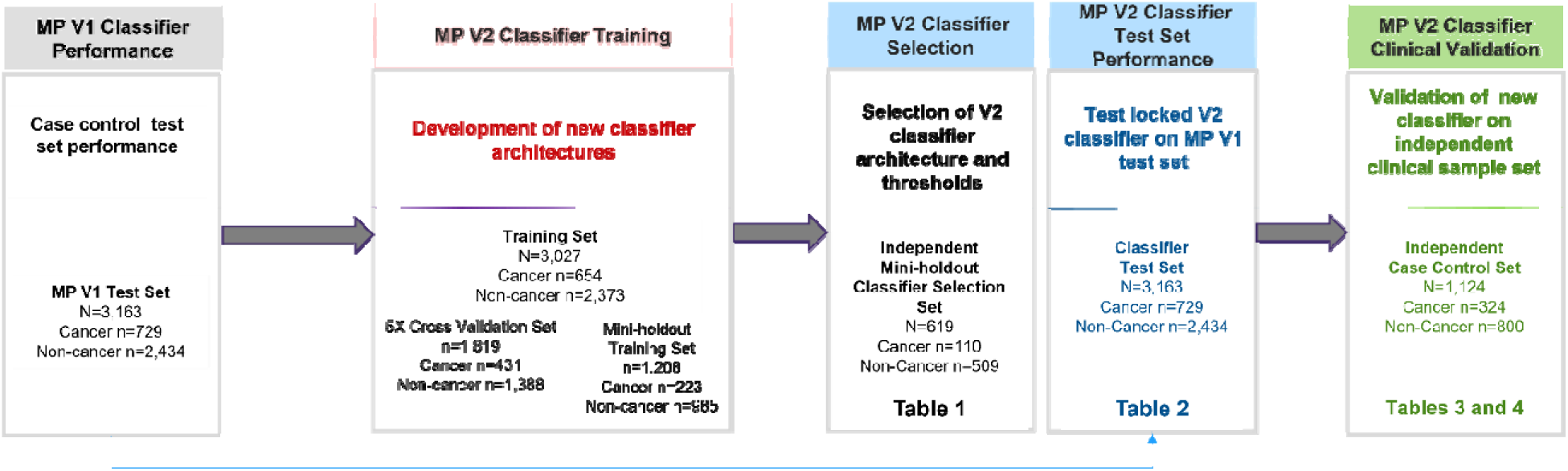
MP V2 classifier development workflow MP, Methylation-Protein.

### Lowering the specificity target and using the MP V1 classifier

Performance is shown in Table 1 and Supplementary Materials Table S1. At a target specificity of ≥98%, the MP V1 classifier resulted in a measured specificity of 99.4% (95% confidence interval [95%CI]: 98.3, 99.8), with an overall sensitivity of 60.9% (95% CI: 51.6, 69.5), and sensitivities of 31.8% (95% CI:16.4, 52.7), 47.7% (95% CI: 33.8, 62.1), 63.6% (95% CI: 43.0, 80.3), 68.2% (95% CI: 47.3, 83.6), 95.7% (95% CI: 79.0, 99.2), and 42.9% (95% CI: 24.5, 63.5) for stages I, I and II combined, II, III, IV, and unknown, respectively for the MHO classifier selection test set (Table 1). When the MP V1 target specificity was lowered to ≥97.0%, the MP V1 classifier (measured specificity of 99.0%; 95% CI: 97.7, 99.6) demonstrated an overall sensitivity of 64.5% (95% CI: 55.3, 72.9), and sensitivities of 36.4% (95% CI: 19.7, 57.0), 52.3% (95% CI: 37.9, 66.2), 68.2% (95% CI: 47.3, 83.6), 77.3% (95% CI: 56.6, 89.9), 95.7% (95% CI: 79.0, 99.2), and 42.9% (95% CI: 24.5, 63.5) for stages I, I and II combined, II, III, IV, and unknown, respectively for the MHO classifier selection test set (Supplementary Materials Table S1).

**Table 1.** Independent mini-holdout classifier selection test set performance comparison.

|  | **MP V1 Classifier**<br>**Target specificity ≥98.0%**<br>(95% CI), n/N | **MP V1 Classifier**<br>**Target specificity ≥97.0%**<br>(95% CI), n/N |  | **MP V2 Classifier**<br>**Target specificity ≥97.0%**<br>(95% CI), n/N |  | **MP V2 Classifier**<br>**Target specificity ≥97.0%**<br>(95% CI), n/N |  |
| --- | --- | --- | --- | --- | --- | --- | --- |
| Measured Specificity | 99.4% (98.3, 99.8), 506/509 | 99.0% (97.7, 99.6), 504/509 |  | 99.0% (97.7, 99.6), 504/509 |  | 99.0% (97.7, 99.6), 504/509 |  |
|  |  | Versus MP V1 classifier<br>@ ≥98% Target Specificity |  | Versus MP V1 classifier<br>@ ≥98% Target Specificity |  | Versus MP V1 classifier<br>@ ≥97% Target Specificity |  |
|  |  | Absolute Increase* | Relative Increase** | Absolute Increase* | Relative Increase** | Absolute Increase* | Relative Increase** |
| Overall Sensitivity | 60.9% (51.6, 69.5), 67/110 | 3.6% | 5.9% | 11.8% | 19.4% | 8.2% | 12.7% |
| Sensitivity by Stage: |  |  |  |  |  |  |  |
| Stage I | 31.8% (16.4, 52.7), 7/22 | 4.6% | 14.5% | 9.1% | 28.6% | 4.5% | 12.4% |
| Stage II | 63.6% (43.0, 80.3), 14/22 | 4.6% | 7.2% | 9.1% | 14.3% | 4.5% | 6.6% |
| Stage III | 68.2% (47.3, 83.6), 15/22 | 9.1% | 13.3% | 18.2% | 26.7% | 9.1% | 11.8% |
| Stage IV | 95.7% (79.0, 99.2), 22/23 | 0.0% | 0.0% | 4.3% | 4.5% | 4.3% | 4.5% |
| Unknown | 42.9% (24.5, 63.5), 9/21 | 0.0% | 0.0% | 19.0% | 44.3% | 19.0% | 44.3% |
| Stages I-II | 47.7% (33.8, 62.1), 21/44 | 4.6% | 9.6% | 9.1% | 19.1% | 4.5% | 8.6% |

\*Absolute Increase=Sensitivity B% - Sensitivity A%. \*\* Relative Increase=Sensitivity B% – Sensitivity A%/Sensitivity A% x 100.

By lowering the MP V1 target specificity by 1% (≥98.0% to ≥97.0%), we observed absolute sensitivity increases of 3.6%, 4.6%, 4.6%, and 4.6% for overall, stage I, II, and stages I and II combined, respectively, in the MHO classifier selection test set (Table 1). These sensitivity increases are equivalent to relative sensitivity increases of 5.9%, 14.5%, 7.2%, and 9.6% for overall, stage I, II, and stages I and II combined, respectively.

### Incorporating improved classifier model architecture and a lower specificity target

The MP V2 classifier at a target specificity of ≥97.0% (measured specificity of 99.0%; 95% CI: 97.7, 99.6) demonstrated an overall sensitivity of 72.7% (95% CI: 63.7, 80.2), and sensitivities of 40.9% (95% CI: 23.3, 61.3), 56.8% (95% CI: 42.2, 70.3), 72.7% (95% CI: 51.8, 86.8), 86.4% (95% CI: 66.7, 95.3), 100.0% (95% CI: 85.7, 100.0), and 61.9% (95% CI: 40.9, 79.2) for stages I, I and II combined, II, III, IV, and unknown, respectively in the MHO classifier selection test set (Supplementary Materials Table S1).

### Evaluating the impact of changes to the specificity target and changes to the model architecture

MP V2 classifier performance (target specificity of ≥97%) was compared to the MP V1 classifier performance (target specificity of ≥98%). Compared to the MP V1 classifier (measured specificity of 99.4%), the MP V2 classifier (measured specificity of 99.0%) resulted in absolute sensitivity increases of 11.8%, 9.1%, 9.1%, and 9.1% for overall, stage I, II, and stages I and II combined, respectively, in the MHO classifier selection test set (Table 1). These absolute sensitivity increases are equivalent to relative sensitivity increases of 19.4%, 28.6%, 14.3%, and 19.1% for overall, stage I, II, and stages I and II combined, respectively (Table 1),

### Isolating the impact of model architecture evolution

To evaluate the performance contributions that may be attributed to improved MP V2 model architecture versus a lowered specificity, MP V2 and MP V1 classifier performances were compared at the same target specificity of ≥97% (measured specificity of 99.0%). Compared to MP V1, MP V2 demonstrated absolute sensitivity increases of 8.2%, 4.5%, 4.5%, and 4.5% for overall, stage I, II, and stages I and II combined, respectively, in the MHO classifier selection test set. These absolute sensitivity increases are equivalent to relative sensitivity increases of 12.7%, 12.4%, 6.6%, and 8.6% for overall, stage I, II, and stages I and II combined, respectively (Table 1). The performance results above suggest that a substantial portion of the observed increases in stage I/II sensitivity may be attributed to MP V2 model architecture improvements.

### MP V1 and V2 Classifier Test Set Performance Comparison

The locked MP V2 classifier was then evaluated using the original ASCEND-2 test development set cohort that was used to evaluate the previously reported MP V1 classifier performance (Figure 1).^15^ At a measured specificity of 97.4%, MP V2 demonstrated improvements across all stage groups compared to MP V1 (specificity 98.5%) (Table 2). Overall sensitivity increased from 50.9% (MP V1) to 57.8% (MP V2), with stage-specific sensitivity increases of 6.0% for stage I, 8.0% for stage II, and 6.9% for stage I–II combined. Relative sensitivity increases were 39.0% for stage I, 21.1% for stage II, and 26.4% for stage I–II combined. Detailed results by stage are shown in Table 2. The differences in absolute sensitivity were statistically significant by McNemar’s test (p < 0.01) for all comparisons except stage IV (p=0.039) and stage unknown (p=0.125). V2 and V1 methylation sub-models demonstrated similar discrimination (AUROC 0.83 and 0.81, respectively) for this dataset. At equivalent targeted specificity (>97.0%), MP V2 classifier showed improvements compared to MP V1 in early-stage sensitivity (33.0% vs 31.0%) and specificity (97.4% vs 97.2%), however these results were not statistically significant.

**Table 2.**
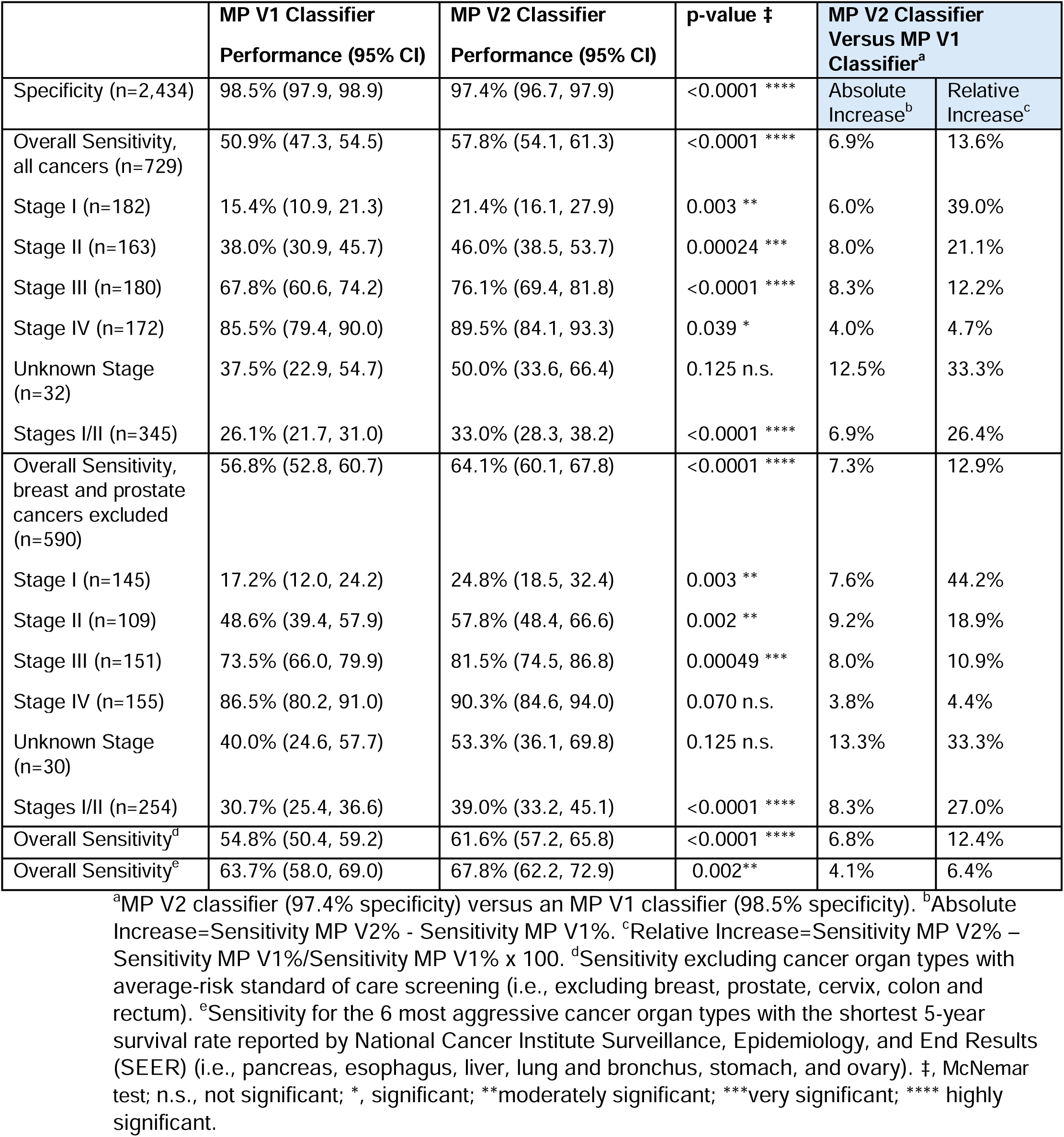
MP V1 and V2 classifier performance in the test set.

|  | MP V1 Classifier<br>Performance (95% CI) | MP V2 Classifier<br>Performance (95% CI) | p-value ‡ | MP V2 Classifier<br>Versus MP V1<br>Classifier<sup>a</sup> |  |
| --- | --- | --- | --- | --- | --- |
| Specificity (n=2,434) | 98.5% (97.9, 98.9) | 97.4% (96.7, 97.9) | <0.0001 **** | Absolute<br>Increase<sup>b</sup> | Relative<br>Increase<sup>c</sup> |
| Overall Sensitivity,<br>all cancers (n=729) | 50.9% (47.3, 54.5) | 57.8% (54.1, 61.3) | <0.0001 **** | 6.9% | 13.6% |
| Stage I (n=182) | 15.4% (10.9, 21.3) | 21.4% (16.1, 27.9) | 0.003 ** | 6.0% | 39.0% |
| Stage II (n=163) | 38.0% (30.9, 45.7) | 46.0% (38.5, 53.7) | 0.00024 *** | 8.0% | 21.1% |
| Stage III (n=180) | 67.8% (60.6, 74.2) | 76.1% (69.4, 81.8) | <0.0001 **** | 8.3% | 12.2% |
| Stage IV (n=172) | 85.5% (79.4, 90.0) | 89.5% (84.1, 93.3) | 0.039 * | 4.0% | 4.7% |
| Unknown Stage<br>(n=32) | 37.5% (22.9, 54.7) | 50.0% (33.6, 66.4) | 0.125 n.s. | 12.5% | 33.3% |
| Stages I/II (n=345) | 26.1% (21.7, 31.0) | 33.0% (28.3, 38.2) | <0.0001 **** | 6.9% | 26.4% |
| Overall Sensitivity,<br>breast and prostate<br>cancers excluded<br>(n=590) | 56.8% (52.8, 60.7) | 64.1% (60.1, 67.8) | <0.0001 **** | 7.3% | 12.9% |
| Stage I (n=145) | 17.2% (12.0, 24.2) | 24.8% (18.5, 32.4) | 0.003 ** | 7.6% | 44.2% |
| Stage II (n=109) | 48.6% (39.4, 57.9) | 57.8% (48.4, 66.6) | 0.002 ** | 9.2% | 18.9% |
| Stage III (n=151) | 73.5% (66.0, 79.9) | 81.5% (74.5, 86.8) | 0.00049 *** | 8.0% | 10.9% |
| Stage IV (n=155) | 86.5% (80.2, 91.0) | 90.3% (84.6, 94.0) | 0.070 n.s. | 3.8% | 4.4% |
| Unknown Stage<br>(n=30) | 40.0% (24.6, 57.7) | 53.3% (36.1, 69.8) | 0.125 n.s. | 13.3% | 33.3% |
| Stages I/II (n=254) | 30.7% (25.4, 36.6) | 39.0% (33.2, 45.1) | <0.0001 **** | 8.3% | 27.0% |
| Overall Sensitivity<sup>d</sup> | 54.8% (50.4, 59.2) | 61.6% (57.2, 65.8) | <0.0001 **** | 6.8% | 12.4% |
| Overall Sensitivity<sup>e</sup> | 63.7% (58.0, 69.0) | 67.8% (62.2, 72.9) | 0.002** | 4.1% | 6.4% |

<sup>a</sup>MP V2 classifier (97.4% specificity) versus an MP V1 classifier (98.5% specificity). <sup>b</sup>Absolute Increase=Sensitivity MP V2% - Sensitivity MP V1%. <sup>c</sup>Relative Increase=Sensitivity MP V2% – Sensitivity MP V1%/Sensitivity MP V1% x 100. <sup>d</sup>Sensitivity excluding cancer organ types with average-risk standard of care screening (i.e., excluding breast, prostate, cervix, colon and rectum). <sup>e</sup>Sensitivity for the 6 most aggressive cancer organ types with the shortest 5-year survival rate reported by National Cancer Institute Surveillance, Epidemiology, and End Results (SEER) (i.e., pancreas, esophagus, liver, lung and bronchus, stomach, and ovary). ‡, McNemar test; n.s., not significant; \*, significant; \*\*moderately significant; \*\*\*very significant; \*\*\*\* highly significant.

MP V1 and MP V2 classifier performance was also compared using the test development set with breast and prostate cancers excluded. At a specificity of 97.4%, MP V2 demonstrated improvements across all stage groups compared to MP V1 (specificity 98.5%) (Table 2). Overall sensitivity increased from 56.8% (MP V1) to 64.1% (MP V2), with stage-specific improvements of 7.6% for stage I, 9.2% for stage II, and 8.3% for stage I–II combined. Relative sensitivity increases were 44.2% for stage I, 18.9% for stage II, and 27.0% for stage I–II combined.

Detailed results by stage are shown in Table 2. The differences in absolute sensitivity were statistically significant by McNemar’s test (p < 0.01) for all comparisons except stage IV (p=0.070) and stage unknown (p=0.125).

Overall sensitivities by organ type for MP V1 (measured 98.5% specificity) and MP V2 (measured 97.4% specificity) classifiers using the test development set ranged from 11.8% for prostate to 80.0% for liver and bile duct (Supplementary Materials Figure S2). MP V1 and MP V2 performance comparisons by organ type and stage are shown in Supplementary Materials Table S2. MP V2 absolute increase in sensitivity was statistically significant for breast (p-value=0.016), bladder (p-value=0.031), and colorectal (p-value=0.002) cancers.

### MP V2 Independent Clinical Validation Set: Participant Characteristics

MP V2 classifier performance was also evaluated in an independent clinical validation (N=1,124; 324 cancer and 800 non-cancer) test set consisting of previously untested samples from the ASCEND-2 study (Figure 1). The clinical validation set consisted of 1,124 subjects with mean age of 64.8 years; 51.2% were female. 83.7% of the clinical validation cohort identified as White, 10.8% Black or African American, and 1.1% Asian; 83.6% were of non-Hispanic/Latino ethnicity. Fifty-three percent were never cigarette smokers, and 50.4% indicated current alcohol consumption. The mean body mass index was 30.0 kg/m^2^, and 56.3% indicated a family history of cancer in a first-degree relative. Cancer and non-cancer participants had similar age, sex, and race/ethnicity, body mass index, and cigarette smoking/alcohol consumption distributions (Table 3). This sample set was selected to more closely resemble the intended use population.

**Table 3.** MP V2 clinical validation set participant demographics and clinical characteristics.

|  | Characteristic | Cancer n (%) | Non-Cancer n (%) | Total N (%) |
| --- | --- | --- | --- | --- |
| **Sex** | Female | 160 (49.4%) | 415 (51.9%) | 575 (51.2%) |
|  | Male | 164 (50.6%) | 385 (48.1%) | 549 (48.8%) |
| **Age (years)** | Mean (SD) | 66.9 (8.2) | 63.7 (8.9) | 64.6 (8.8) |
|  | (Min, Max) | (50, 84) | (50, 84) | (50, 84) |
| **Race** | White | 284 (87.7%) | 657 (82.1%) | 941 (83.7%) |
|  | Black or African American | 30 (9.3%) | 91 (11.4%) | 121 (10.8%) |
|  | Asian | 2 (0.6%) | 10 (1.3%) | 12 (1.1%) |
|  | American Indian or Alaska Native | 2 (0.6%) | 6 (0.8%) | 8 (0.7%) |
|  | Native Hawaiian or Other Pacific Islander | 1 (0.3%) | 1 (0.1%) | 2 (0.2%) |
|  | More than one race | 1 (0.3%) | 6 (0.8%) | 7 (0.6%) |
|  | Unknown | 4 (1.2%) | 29 (3.6%) | 33 (2.9%) |
| **Ethnicity** | Hispanic/Latino | 11 (3.4%) | 155 (19.4%) | 166 (14.8%) |
|  | Not Hispanic/Latino | 302 (93.2%) | 636 (79.5%) | 938 (83.6%) |
|  | Unknown | 11 (3.4%) | 7 (0.9%) | 18 (1.6%) |
|  | Missing | 0 | 2 | 2 |
| **BMI (kg/m<sup>2</sup>)** | Mean (SD) | 29.4 (7.1) | 30.2 (6.7) | 30.0 (6.8) |
|  | (Min, Max) | (14.2, 61) | (16.4, 66.4) | (14.2, 66.4) |
|  | Missing | 1 | 0 | 1 |
| **Cigarette Smoking History** | Current | 50 (15.5%) | 118 (14.8%) | 168 (15.0%) |
|  | Former | 141 (43.8%) | 215 (26.9%) | 356 (31.7%) |
|  | Never | 131 (40.7%) | 467 (58.4%) | 598 (53.3%) |
|  | Missing | 2 | 0 | 2 |
| **Alcohol Consumption History** | Current | 153 (47.5%) | 411 (51.6%) | 564 (50.4%) |
|  | Former | 55 (17.1%) | 95 (11.9%) | 150 (13.4%) |
|  | Never | 114 (35.4%) | 291 (36.5%) | 405 (36.2%) |
|  | Missing | 2 | 3 | 5 |
| **Family History of Cancer in 1<sup>st</sup> Degree Relative** | No | 105 (32.4%) | 363 (45.4%) | 468 (41.7%) |
|  | Yes | 208 (64.2%) | 424 (53.1%) | 632 (56.3%) |
|  | Unknown | 11 (3.4%) | 12 (1.5%) | 23 (2.0%) |
|  | Missing | 0 | 1 | 1 |

BMI, body mass index kg/m<sup>2</sup>

### MP V2 Classifier Independent Clinical Validation Set Performance

Using the independent clinical validation set, the MP V2 classifier, at a measured specificity of 97.4% (95% CI: 96.0, 98.3), showed an overall sensitivity of 41.4% (95% CI: 36.1, 46.8), and sensitivities of 16.0% (95% CI: 10.3, 24.2), 22.8% (95% CI: 17.4, 29.2), 31.3% (95% CI: 22.4, 41.9), 52.9% (95% CI: 41.3, 64.1), and 83.1% (95% CI: 72.2, 90.3) for stages I, I and II combined, II, III, and IV, respectively (Table 4). Overall sensitivity excluding cancer organ types with average-risk, single-cancer screening (i.e. excluding breast, prostate, cervix, colon and rectum) was 52.4% (95% CI: 43.7, 60.9) and 67.9% (95% CI: 58.5, 76.0) for the six most aggressive cancer organ types with the shortest 5-year survival rate (i.e. pancreas, esophagus, liver, lung and bronchus, stomach, and ovary).^23^ With breast and prostate cancers excluded, the MP V2 classifier demonstrated an overall sensitivity of 55.6% (95% CI: 49.0, 62.0), and sensitivities of 26.8% (95% CI: 17.0, 39.6), 34.8% (95% CI: 26.6, 44.0), 42.9% (95% CI: 30.8, 55.9), 63.6% (95% CI: 50.4, 75.1), and 89.3% (95% CI: 78.5, 95.0) for stages I, I and II combined, II, III, and IV, respectively (Table 4). The MP V2 classifier overall sensitivities by organ type ranged from 3.9% (prostate) to 100.0% (ovary, small intestine, anus, vulva, and testis) (Supplementary Materials Table S4) in the clinical validation set. Table S4 also shows the performance by stage.

**Table 4.** MP V2 classifier validation set performance.

| (n/N) | Performance | 95% CI |
| --- | --- | --- |
| Specificity (779/800) | 97.4% | (96.0, 98.3) |
| Overall Sensitivity, all cancers (134/324) | 41.4% | (36.1, 46.8) |
| Stage I (17/106) | 16.0% | (10.3, 24.2) |
| Stage II (26/83) | 31.3% | (22.4, 41.9) |
| Stage III (37/70) | 52.9% | (41.3, 64.1) |
| Stage IV (54/65) | 83.1% | (72.2, 90.3) |
| Stages I-II (43/189) | 22.8% | (17.4, 29.2) |
| Overall Sensitivity, breast and prostate cancers excluded (124/233) | 55.6% | (49.0%, 62.0) |
| Stage I (15/56) | 26.8% | (17.0, 39.6) |
| Stage II (24/56) | 42.9% | (30.8, 55.9) |
| Stage III (35/55) | 63.6% | (50.4, 75.1) |
| Stage IV (50/56) | 89.3% | (78.5, 95.0) |
| Stages I-II (39/112) | 34.8% | (26.6%, 44.0) |
| Overall Sensitivity* (66/126) | 52.4% | (43.7, 60.9) |
| Overall Sensitivity** (72/106) | 67.9% | (58.5, 76.0) |

\*Sensitivity excluding cancer organ types with average-risk standard of care screening (i.e., excluding breast, prostate, cervix, colon and rectum). \*\*Sensitivity for the 6 most aggressive cancer organ types with the shortest 5-year survival rate reported by National Cancer Institute Surveillance, Epidemiology, and End Results (SEER) (i.e., pancreas, esophagus, liver, lung and bronchus, stomach, and ovary).

### Cancer Subtype Analysis

The MP test is based on sets of shared methylation and protein biomarkers present in many different tumor types; these biomarkers have the potential to detect a broad range of cancers from a blood specimen. The case report forms obtained from 635 subjects with cancer (421 from the MP V2 test development set, 80 from the MHO test set, and 134 from the independent clinical validation test set) were assessed to determine the tumor types and histological subtypes detected. Over fifty distinct cancer subtypes were identified. Among those, 17 cancer subtypes were identified based on single cases (n=1). Cancer subtype staging was not determined. The cancer subtypes detected are listed in Supplementary Materials Table S5.

## Discussion

MCED testing has the potential to shift population-wide cancer screening from single-cancer screening of a handful of cancers to many cancers through expanded multi-cancer screening using a single blood specimen. MCED tests are typically designed with high specificity targets to limit FP results and the associated harms associated with futile diagnostic procedures.^24^ Classifier development is challenging as it requires balancing case-control derived sensitivity/specificity measurements to project real world-clinical performance. We therefore attempted to develop a refined classifier that is more generalizable to cohort differences and target a lower target specificity in a case-control setting. In this study, we first identified and selected a new model architecture and then adjusted the specificity target to evaluate performance in a classifier selection set, a previously described test set, and finally in an independent validation set more closely representative of the intended use population. We found that incorporating MP V2 model architecture improvements and lowering the specificity target by one percent resulted in a 6.9% absolute (26.4% relative) sensitivity increase for early-stage cancers (stage I and II combined) compared to the MP V1 classifier. A detailed analysis of the MHO classifier selection set showed that the observed performance improvements are due to both the lower case-control target specificity and the new model architecture. The MP V2 classifier will be utilized in a real-world setting to further validate the sensitivity and specificity results.

False positive results are of concern for both single cancer and MCED tests. However, since MCED tests interrogate cancer signals originating from multiple organ sites, FP MCED results could trigger diagnostic procedures that may impose physical and psychological burdens on patients to definitively rule in or rule out cancer. Specificities of ≥98.9% have been reported for the prospective PATHFINDER, PATHFINDER-2, and DETECT-A studies, resulting in FP rates of ≤2%,^17, 18, 19^ much lower than FP rates reported for guideline-recommended single-cancer screening tests.^12^ An invasive procedure (e.g., surgery) was required to resolve FP results in just 2%, 0.4%, and 3% of cases, respectively in the aforementioned studies.^17, 18, 19^ Efficient imaging-based diagnostic resolution strategies may further reduce the potential risks associated with the workup for FP MCED test results.^25^

This study has some limitations. Of the projected two million new cancer diagnoses in 2026,^1^ it is estimated that approximately 88% (almost 1.8 million) will occur in individuals aged 50 years and older.^26^ In addition, it is likely that the prevalence of undiagnosed cancers harbored by apparently healthy individuals at any point in time is higher as many cancers have pre-clinical dwell times that extend beyond one year.^27^ The controls used to develop and test the MP V1 classifier included age-matched individuals without suspicion of cancer. While free from suspicion of cancer at the time of enrollment, controls were not required to have completed all recommended cancer screening tests, were not subject to any procedures to rule out the presence of an occult cancer and participants were not followed after contributing a blood sample to confirm the absence of cancer that might emerge in subsequent months. Therefore, the control group likely included individuals with hidden, undiagnosed cancers. Thus, targeting a specificity of ≥98.5% in the MP V1 classifier development case-control study may have been conservative, given that measured specificity was likely slightly underestimated by undiagnosed cancers in the control group, potentially sacrificing sensitivity.

Although the ASCEND-2 samples used in this study were prospectively collected from more than 200 sites across the US and Europe and included a diverse participant cohort broadly representative of the U.S. population, the analyses were performed with cases having tumor type representation that is close to an intended use population but with equal stage distributions across several cancer types. Therefore, the ASCEND -2 case-control performance results might be different than real-world test performance. Both the MP V1 and MP V2 classifiers were evaluated using the same test set design. When MP V2 classifier sensitivity was assessed in a clinical validation cohort that more closely reflected SEER incidence, the observed overall sensitivity was 41.4%. MCED test performance must be interpreted with considerable caution, as differences in cancer types/stages affect performance estimates. Previous studies have reported significant differences in performance between case-control and prospective interventional studies,^17–19, 21, 22^ even for the same assay in different prospective studies.^18, 19^ While improvements in performance due to test development can be assessed in case-control studies, as reported here, performance in the screening setting is determined in prospective studies in the intended-use population. Therefore, performance comparisons across studies should account for differences in design, cancer type mix, and stage distribution, and simple cross-study comparisons should be avoided.

In summary, as part of an ongoing MCED test development process, we trained and tested a refined MCED classifier, MP V2. We adjusted a pre-specified case-control specificity target from ≥98% to ≥97% to optimize early-stage (stages I and II) cancer detection which should result in maintained or increased specificity in a real-world setting.^17, 18, 21, 22^ The locked MP V2 classifier was then evaluated in a clinical validation cohort selected to more closely resemble a real-world population. Prospective, real-world evaluation of the MP V2 classifier will be needed to determine if we achieved the optimal balance between sensitivity and specificity.

## Methods

### MP V1 Classifier

A prospectively collected, case-control study, ASCEND-2, was the primary sample source for the development of the MP V1 classifier.^15^ MP V1 classifier development included participants from multiple sites across the United States and within Europe. The MP V1 classifier development cohort included male and female subjects ≥50 years old with known cancer and controls without suspicion of cancer. Cases were recruited immediately following cancer diagnosis and before treatment initiation or, in the case of cancer types that are diagnosed with excisional biopsies, immediately prior to the diagnostic and therapeutic procedure. All collection sites had Institutional Review Board approval, and all eligible subjects provided written informed consent and were assessed for study participation eligibility. Blood samples were collected in LBgard^®^ tubes at the study sites, shipped to centralized laboratories, and processed into plasma and buffy aliquots. Protein and methylation assays were assessed using separate plasma aliquots.

### Protein Analysis

Plasma protein concentrations were measured using commercially available assays for 6 proteins on a Roche cobas e801 instrument (Roche Diagnostics, Indianapolis, IN) per manufacturer’s recommendations.^28^ The cobas e801 analyzer uses electrochemiluminescence (ECL)-based immunoassays to measure protein concentrations.

### Methylated DNA Marker (MDM) Analysis

Methylated DNA markers were measured as previously described.^28^ Briefly, circulating tumor (ctDNA) was extracted from 6 mL plasma followed by bisulfite conversion on a Hamilton Microlab STARlet (Hamilton Robotics, Reno, NV) using reagents manufactured by Exact Sciences Corporation (Madison, WI). Bisulfite converted ctDNA was then analyzed using Target Enrichment Long-probe Quantitative Amplified Signal (TELQAS^TM^) chemistry. For the initial target enrichment, converted ctDNA was transferred to a 96-well polymerase chain reaction (PCR) plate and subjected to multiplex PCR to amplify the MDMs and a reference methylation marker. The PCR products were diluted 10-fold and evaluated in triplex or biplex assays using Long-probe Quantitative Amplified Signal (LQAS^TM^) on a Quantstudio 5 Dx Real-Time PCR Instrument (Life Technologies Corporation, Grand Island, NY). TELQAS combines amplification using real-time PCR and allele-specific detection of methylated target DNA through an invasive cleavage assay.

### MP V2 Classifier Training

The overall MP V2 classifier development workflow is shown in Figure 1. The MP V2 classifier candidates were trained using the same methylation and protein data set previously used for the training of MP V1^15^. The total training set was comprised of 3,027 samples (654 cancer and 2,373 non-cancer), that was partitioned into two sub-groups for 5-fold cross validation (N=1,819; 431 cancer and 1,388 non-cancer) and MHO testing (N=1,208; 223 cancer and 985 non-cancer). The MP V2 training set was used to investigate multiple alternative classifier architectures, feature engineering, and transformation approaches as well as to perform tuning of specificity thresholds. A target specificity of ≥97.0% was used for MP V2 classifier training. Only samples that passed pre-specified methylation and protein assay quality control criteria were included in the training process; these criteria were identical for the MP V1 and MP V2 training processes. Methylation and protein classifiers were trained individually, and the individual classifier results were combined using a logical OR overarching classifier to generate the final (positive or negative) call. The result was deemed positive when either the methylation or protein assay provided a cancer signal detected call.

### MP V2 Classifier Model Selection

After multiple potential classifiers were developed and evaluated as part of training, locked candidate models were compared during a classifier selection step using newly generated methylation and protein data from an independent MHO classifier selection test set (MHO; n=619; n=110 Cancers; n=509 non-cancers). The samples were selected from a previously untested cohort of subjects of the ASCEND-2 study cohort.^15^ The performance of the MP V2 candidate classifiers was compared to the MP V1 classifier at two different target specificities (original target of ≥98.0% and updated target of ≥97.0%). A Feature Weighted Naive Bayes (FWNB) classifier was selected as it showed improved performance compared to both MP V1 and other candidate MP V2 model architectures. This model was locked for additional tuning and then subjected to additional testing.

### MP V2 Classifier Evaluation

To enable direct comparison of V2 to V1 MP classifiers, the locked MP V2 classifier was tested using the original data set used for the performance characterization of the MP V1 classifier.^15^ This test set included (n=3,163; 729 cancers and 2,434 non-cancers) and was described above.

The MP V2 performance assessment at the adjusted specificity target (≥97%) was compared to the MP V1 classifier at the original (≥98%) specificity target.

### MP V2 Classifier Clinical Validation

The MP V2 classifier performance was further evaluated in an independent clinical validation (N=1,124; 324 cancer and 800 non-cancer) test set. The samples for this set were selected from previously untested subjects of the ASCEND-2 study cohort.^15^ The clinical validation test set cancer cohort included nineteen organ sites (prostate (n=51), breast (n=50), lung and bronchus (n=57), colon and rectal (n=38), kidney (n=19), uterus (n=17), head and neck (squamous cell; n=17), pancreas (n=17), bladder and urinary (n=12), liver (n=12), thyroid (n=5), stomach (n=7), ovary (n=7), esophagus (n=6), small intestine (n=3), cervix uteri (n=2), anus (n=2), vulva (n=1) and testis (n=1)). The stage distributions for these organ tumor sites are shown in Supplementary Materials Table S3.

Cancer samples were selected to achieve balanced representation across stages I-IV (excluding breast and prostate cancer) for estimation of stage-specific performance. Within stage, the distribution of seventeen organ sites (excluding breast and prostate) was chosen to reflect 2021 incidence reported by the SEER database.^23^ For breast and prostate cancer, we selected fifty samples each, ensuring the stage distribution reflected 2021 SEER incidence.

Non-cancer samples were chosen so that the distribution of age groups, sex, and race mirrored the demographics reported in the 2021 Census, to more closely represent the intended use population in a real-world setting.^29^ The clinical validation participant demographics and clinical characteristics are shown in Table 3.

### Statistical Analysis

Sensitivity and specificity were estimated overall and stratified by cancer stage. Sensitivity was defined as the proportion of participants with disease who tested positive, and specificity as the proportion of participants without disease who tested negative. Two-sided 95% confidence intervals were calculated using the Wilson score method. Comparisons of sensitivity and specificity between classifiers were performed using McNemar’s test for paired binary outcomes, applied within each stage group and for the overall cohort. Statistical significance was assessed using two-sided tests with a threshold of p < 0.05. SAS software (https://www.sas.com) was used for CV analysis, Python SciPy and statsmodels (https://www.statsmodels.org), scikit-learn (https://www.scikit-learn.org) was used for model development.

### Cancer Subtype Analysis

Case report forms were assessed by three independent physicians to identify biopsy and histopathology variables for each detected tumor type across the cohorts. The cancers were classified into distinct histological types and subtypes based on cancer site, morphology, and international classification of diseases for oncology (ICD-O) codes. Certain cancer types were designated as unique due to differences in histology, pathogenesis, risk factors, and clinical management, reflecting their recognition as separate entities in clinical practice. Limited cancer type, histology subtype, and clinical information were available for this analysis, and certain histological subtypes were not classified as distinct cancer types. With limited numbers of cases that represent many cancer subtypes, these data should not be used to estimate real world test performance by cancer subtypes.

## Data Availability

The data generated in this study is available upon request from the corresponding author.

## Acknowledgements

The authors would like to thank Michelle Beidelschies for leadership and feedback.

Carolyn Hall and Feyza Sancar (Exact Sciences Corporation, Madison, WI) provided manuscript writing and editorial support. The authors would also like to thank the patients who generously participated in the original sample collection studies and the principal investigators and institutions who oversaw their enrollment. This study was funded by Exact Sciences Corporation.

## Author Contributions

Conceptualization: Frank Diehl Methodology: Vladimir Gainullin, Frank Diehl

Formal Analysis: Vladimir Gainullin, Melissa Gray, Madhav Kumar, Stephen Luebker, Amy Lehman, Omair Choudhry, Julia Roberta, Darl Flake, Avinash Shanmugam, Kevin Cortes, Emily Chang, Philip J. Uren

Writing-Original Draft: Vladimir Gainullin, Frank Diehl, Tomasz M. Beer

Writing-Review and Editing: Vladimir Gainullin, Melissa Gray, Madhav Kumar, Stephen Luebker, Amy Lehman, Omair Choudhry, Julia Roberta, Darl Flake, Avinash Shanmugam, Kevin Cortes, Emily Chang, Philip J. Uren, Amin Mazloom, Jorge Garces, Tomasz M. Beer, Frank Diehl

Project Administration: Amin Mazloom, Jorge Garces, Frank Diehl

## Conflicts of Interest

**V.G.G.**, **M.G.**, **M.K., S.L., A.M.L., O.A.C., J.R., D.D.F. II, A.S., K.C., E.C., P.J.U., J.G., A.M., T.M.B.**, and **F.D.** are employees at Exact Sciences Corporation and declare stock ownership at Exact Sciences Corporation. **A.M., J.G.**, **T.M.B.**, and **F.D.** hold leadership roles at Exact Sciences Corporation. **P.J.U.** declares stock ownership at Biora Therapeutics outside this work. **T.M.B.** declares stock ownership at Osteologic and Osheru, and a consulting/advisory role at AstraZeneca outside this work. **G.A.S.** declares advisory board service at Candel Therapeutics, consulting fees from Freenome and Biodesix, and research funding (Institutional) from NIH (RO1 screening in frail populations), Nucleix, Inc, Biodesix, Delfi Diagnostics, and Freenome outside this work. **G.A.S.** serves on the American Cancer Society roundtable steering committee outside this work. **D.W.C.** declares no conflicts of interest. **R.W.G.** declares speaker’s bureau service at Bayer and Johnson & Johnson, research funding from Francis Medical, MDX Health, Levee, and Dendreon; and travel reimbursement from Francis Medical outside this work.

## Notes

### Competing Interest Statement

Vladimir Gainullin, Melissa Gray, Madhav Kumar, Stephen Luebker, Amy M. Lehman, Omair A. Choudhry, Julia Roberta, Darl D. Flake II, Avinash Shanmugam, Kevin Cortes, Emily Chang, Philip J. Uren, Jorge Garces, Amin Mazloom, Tomasz M. Beer, and Frank Diehl are employees at Exact Sciences Corporation and declare stock ownership at Exact Sciences Corporation. Amin Mazloom, Jorge Garces, Tomasz M. Beer, and Frank Diehl hold leadership roles at Exact Sciences Corporation. Philip J. Uren declares stock ownership at Biora Therapeutics outside this work. Tomasz M. Beer declares stock ownership at Osteologic and Osheru, and a consulting/advisory role at AstraZeneca outside this work. Gerard A. Silvestri declares advisory board service at Candel Therapeutics, consulting fees from Freenome and Biodesix, and research funding (Institutional) from NIH (RO1 screening in frail populations), Nucleix, Inc, Biodesix, Delfi Diagnostics, and Freenome outside this work. Gerard A. Silvestri serves on the American Cancer Society roundtable steering committee outside this work. David W. Chesla declares no conflicts of interest. Robert W. Given declares speakers bureau service at Bayer and Johnson & Johnson, research funding from Francis Medical, MDX Health, Levee, and Dendreon; and travel reimbursement from Francis Medical outside this work.

### Author Declarations

All collection sites had Institutional Review Board approval, and all eligible subjects provided written informed consent and were assessed for study participation eligibility. The IRB Committees of Eastern Connecticut Hematology and Oncology, Mid Florida Cancer Centers Orange City, Joliet Oncology Hematology Associates (JOHA), Adult Pediatric Urology & Urogynecology Omaha, Nebraska, SSM Health Dean Medical Group Madison, The Urology Center of Colorado Denver, The Minniti Center for Hematology and Oncology, Cancer Centers of Southwest Oklahoma Lawton, Lewis Hall Singletary Archbold Cancer Center, MUSC College of Medicine, Meritus Center for Clinical Research, St. Michael Oncology Clinic Cowhorn Creek Research Department, Torrance Memorial Hunt Cancer Institute, Pasadena Center for Medical Research, LLC, Finlay Medical Research Corp West Palm Beach, Delta Research Monroe, NextPhase Research Alliance Cano Health, Allina Health Cancer Institute Minneapolis, New Jersey Cancer Care, PA Belleville, American Research Institute, Inc, Upstate Lung and Critical Care Specialists Spartanburg, Upstate Lung and Critical Care Specialists Union, MercyHealth Hospital and Trauma Center Janesville, Javon Bea Hospital Rockton, Urology South Clinical Campus,1960 Physicians Associates Cypress Creek Location, Gabrail Cancer & Research Center, Town and Country Internal Medicine, Coastal Pulmonary & Critical Care, The Blue Ridge Institute for Medical Research (BRIMR), Mercy Family Clinic & Wellness Center, Tilda Research, Sarasota Memorial Hospital, Biopharma Informatic Los Angeles, CA Campus, Ralph H. Johnson Department of Veterans Affairs Medical Center, Velocity Clinical Research Columbia, Upstate Lung and Critical Care Specialists Greenville, Velocity Clinical Research Anderson, UW Health Carbone Cancer Center, MultiCare Rockwood Clinic Cheney, Urology of Virginia Virginia Beach, Premier Oncology Consultants Memorial Location, Martin O'Neil Cancer Center, Mayo Clinic Rochester, Athens Comprehensive Cancer Center, Corewell Health Grand Rapids Hospitals Butterworth, The Oncology Institute of Hope & Innovation, Virginia Cancer Institute West End, Mercy Clinic Oncology and Hematology Joplin, Mercy Research David C. Pratt Cancer Center, Mercy Clinic Cancer and Hematology Chub O'Reilly Cancer Center, Mercy Clinic Oncology Fort Smith, David M. Sindelar Cancer Center, Mercy Clinic Oncology and Hematology Oklahoma City South, Spartanburg Medical Center, Baptist Health Medical Group Hematology & Oncology at Baptist Health Floyd, Baptist Health Lexington, Baptist Health Louisville, Baptist Health Paducah, Baptist Health Corbin, Baptist Health Hardin Elizabethtown, Fort Wayne South Office, Upstate Lung and Critical Care Specialists Gaffney, Durham VA Medical Center, MD Anderson Cancer Center Gastroenterology, Hepatology and Nutrition Department, Jackson Siegelbaum Gastroenterology East Shore, MyCare Medical McAllen, Jamaica Hospital Medical Center, Samaritan Pastega Regional Cancer Center, MultiCare Regional Cancer Center Tacoma, SMIL Southwest Medical Imaging Research, PRX Research, Idaho Urologic Institute Meridian, The Jackson Clinic Baptist Campus, East View Medical Research, LLC, GastroMed Miami / Coral Gables, Louisiana Research Center, LLC, La Crosse Hospital, Alabama Oncology, Albert Lea Hospital and Clinic, Mankato Hospital and Clinic, Women & Infants Hospital, Wichita Urology, AMPM Research Clinic, Norton Cancer Institute Resource Center Downtown, Knight Cancer Research Building, Eau Claire Hospital and Clinic, Cleveland Clinic Foundation, Polly and Bill Van Dyke Cancer Center Ascension Columbia St. Mary's Hospital Milwaukee Campus, Utah Hematology Oncology South Ogden, Virginia Urology Stony Point, Brooklyn VA Medical Center, Eastchester Cancer Center, Lucy Curci Cancer Center, Mayo Clinic Arizona Office of Clinical Research, Baylor Scott & White Medical Center Temple, American Research Institute, Inc, Upstate Lung and Critical Care Specialists Union, Upstate Lung and Critical Care Specialists Gaffney, Velocity Clinical Research Columbia, Velocity Clinical Research Anderson, Upstate Lung and Critical Care Specialists Spartanburg, Omar A. Gomez, MD, PA, Elligo Health Research, Laguna Clinical Research Associates Laredo Laguna, HealthStar Physicians, WestGlen Gastrointestinal Consultants, Premier Medical Center Toluca Lake, Gilbert Center for Family Medicine, Sugar Lakes Family Practice, Vasantha Pai, MD Gastroenterologist, Digestive and Liver Disease Center of San Antonio, PLLC, Protenium Clinical Research, San Joaquin General Hospital Gastroenterology Clinic, Valley Cancer Associates, NY Family Docs, KMC Dermatology Topeka, Georgia Skin & Cancer Clinic Savannah Office, Boston Clinical Trials, Skylight Health Research North Academy Boulevard, Skylight Health Research Cambridge Street, Aurora Urgent Care, Oviedo Medical Research, IMA Clinical Research Chicago, IMA Clinical Research Manhattan, IMA Clinical Research St. Louis, IMA Clinical Research St. Petersburg, University Health Truman Medical Center, BTC of New Bedford, Elligo Clinical Research Center, Boston Clinical Trials, Gilbert Center for Family Medicine, Georgia Skin & Cancer Clinic Savannah Office, IMA Clinical Research Chicago, IMA Clinical Research Manhattan, IMA Clinical Research St. Louis, IMA Clinical Research St. Petersburg, KMC Dermatology Topeka, Laguna Clinical Research Associates Laredo Laguna, Premier Medical Center Toluca Lake, Oviedo Medical Research, Skylight Health Research Cambridge Street, Skylight Health Research North Academy Boulevard, Sugar Lakes Family Practice, Valley Cancer Associates, Vasantha Pai, MD Gastroenterologist, Valley OB Gyn Clinic Saginaw, MI, Texas Oncology Arlington Cancer Center North, Compass Oncology Vancouver Cancer Center, Texas Oncology Medical City Dallas, Prisma Health Cancer Institute Grove Commons, Texas Oncology Denison Cancer Center, Texas Oncology Amarillo Cancer Center, Texas Oncology McAllen, Shenandoah Oncology, Texas Oncology Bedford, Texas Oncology The Woodlands, Texas Oncology McKinney, Texas Oncology Plano West, Texas Oncology Denton, Texas Oncology Presbyterian Cancer Center Dallas, Mamie McFaddin Ward Cancer Center, Texas Oncology San, Antonio Northeast, Rocky Mountain Cancer Centers Aurora, Woodlands Medical Specialists, Consultants In Medical Oncology and Hematology Broomall, Texas Oncology Tyler, Texas Oncology Deke Slayton Cancer Center, Texas Oncology Fort Worth Cancer Center, Willamette Valley Cancer Institute and Research Center Eugene, Texas Oncology Methodist Dallas Cancer Center, Illinois Cancer Specialists Arlington Heights, Texas Oncology Plano East, Texas Oncology Flower Mound, Texas Oncology Sugar Land, Abington Hematology Oncology Associates, Texas Oncology Methodist Charlton Cancer Center, Texas Oncology The Woodlands, Des Moines Oncology Research Association, Gastroenterologisch onkologische Praxisklinik im Handelhauskarree, Cancer Center Esslingen, Freeman Hospital, Royal Free Hospital, Royal Blackburn Teaching Hospital, Royal Devon and Exeter Hosptal (Wonford), Royal Preston Hospital, Guy's Hospital, Blackpool Victoria Hospital, CWZ Nijmegen, Gelre Ziekenhuis Apeldoorn, Accellacare Coventry, and EB Medical Research gave ethical approval for this work.

### Summary of Updates

Supplementary Table S5 content has been revised.

