## Supplementary material for "Performance of an Optimized Methylation-Protein Multi-Cancer Early Detection (MCED) Test Classifier": MedRxiv Supplementary Materials_3-19-2026.docx

*Former Exact Sciences employee

**Corresponding Author

Frank Diehl, PhD

Senior Vice President, Research & Development, Multi-cancer Early Detection

Exact Sciences Corporation

11085 N. Torrey Pines Rd.

La Jolla, CA 92037

Table S1. MP V1 and MP V2 performance in the independent mini-holdout classifier selection test set

|  | **MP V1 Classifier (95% CI)**  **Target specificity ≥97%** | **MP V2 Classifier** **(95% CI)**  **Target specificity ≥97%** |
| --- | --- | --- |
| Measured Specificity | 99.0% (97.7-99.6%) 504/509 | 99.0% (97.7-99.6%), 504/509 |
| Overall Sensitivity | 64.5% (55.3-72.9%) | 72.7% (63.7-80.2%) |
| Sensitivity by Stage: |  |  |
| Stage I (N=22) | 36.4% (19.7-57.0%) | 40.9% (23.3-61.3%) |
| Stage II (N=22) | 68.2% (47.3-83.6%) | 72.7% (51.8-86.8%) |
| Stage III (N=22) | 77.3% (56.6-89.9%) | 86.4% (66.7-95.3%) |
| Stage IV (N=23) | 95.7% (79.0-99.2%) | 100.0% (85.7-100.0%) |
| Unknown (N=21) | 42.9% (24.5-63.5%) | 61.9% (40.9-79.2%) |
| Stages I-II (N=44) | 52.3% (37.9-66.2%) | 56.8% (42.2-70.3%) |

MP, Methylation-Protein.

Table S2. test set performance comparison between MP V1 and V2 by organ type and stage

| Organ Type | Stage, (n) | MP V1 Classifier* | MP V2 Classifier** |
| --- | --- | --- | --- |
| Lung and Bronchus | Stage I (46)  Stage II (23)  Stage III (48)  Stage IV (39)  Unknown (3) | 13% (6.1%, 25.7%)  52.2% (33.0%, 70.8%)  77.1% (63.5%, 86.7%)  92.3% (79.7%, 97.3%)  100.0% (43.9%, 100%) | 19.6% (10.7%, 33.2%)  52.2% (33.0%, 70.8%)  83.3% (70.4%, 91.3%)  92.3% (79.7%, 97.3%)  100.0% (43.9%, 100.0%) |
| Colon and Rectum | Stage I (13)  Stage II (18)  Stage III (25)  Stage IV (27)  Unknown (5) | 15.4% (4.3%, 42.2%)  50.0% (29%, 71.0%)  76.0% (56.6%, 88.5%)  92.6% (76.6%, 97.9%)  40.0% (11.8%, 76.9%) | 23.1% (8.2%, 50.3%)  61.1% (38.6%, 79.7%)  88.0% (70.0%, 95.8%)  96.3% (81.7%, 99.3%)  100.0% (56.6%,100.0%) |
| Breast | Stage I (24)  Stage II (30)  Stage III (23)  Stage IV (9)  Unknown (2) | 8.3% (2.3%, 25.8%)  30.0% (16.7%, 47.9%)  47.8% (29.2%, 67%)  88.9% (56.5%, 98%)  0.0% (0.0%, 65.8%) | 8.3% (2.3%, 25.8%)  40.0% (24.6%, 57.7%)  60.9% (40.8%, 77.8%)  100.0% (70.1%, 100.0%)  0.0% (0.0%, 65.8%) |
| Prostate | Stage I (13)  Stage II (24)  Stage III (6)  Stage IV (8)  Unknown (0) | 7.7% (1.4%, 33.3%)  0.0% (0.0%, 13.8%)  0.0% (0.0%, 39.0%)  62.5% (30.6%, 86.3%)  - | 7.7% (1.4%, 33.3%)  0.0% (0.0%, 13.8%)  0.0% (0.0%, 39.0%)  62.5% (30.6%, 86.3%)  - |
| Uterus | Stage I (20)  Stage II (4)  Stage III (7)  Stage IV (6)  Unknown (2) | 15.0% (5.2%, 36%)  25.0% (4.6%, 69.9%)  57.1% (25%, 84.2%)  83.3% (43.6%, 97.0%)  0.0% (0.0%, 65.8%) | 15.0% (5.2%, 36%)  25.0% (4.6%, 69.9%)  71.4% (35.9%, 91.8%)  83.3% (43.6%, 97.0%)  50.0% (9.5%, 90.5%) |
| Pancreas | Stage I (7)  Stage II (8)  Stage III (7)  Stage IV (14)  Unknown (1) | 42.9% (15.8%, 75.0%)  50.0% (21.5%, 78.5%)  100.0% (64.6%, 100.0%)  92.9% (68.5%, 98.7%)  0.0% (0.0%, 79.3%) | 42.9% (15.8%, 75.0%)  75.0% (40.9%, 92.9%)  100.0% (64.6%, 100.0%)  92.9% (68.5%, 98.7%)  0.0% (0.0%, 79.3%) |
| Head and Neck | Stage I (10)  Stage II (5)  Stage III (11)  Stage IV (10)  Unknown (0) | 40.0% (16.8%, 68.7%)  40.0% (11.8%, 76.9%)  54.5% (28%, 78.7%)  80.0% (49%, 94.3%)  - | 60.0% (31.3%, 83.2%)  60.0% (23.1%, 88.2%)  72.7% (43.4%, 90.3%)  80.0% (49.0%, 94.3%)  - |
| Kidney | Stage I (9)  Stage II (4)  Stage III (4)  Stage IV (11)  Unknown (4) | 0.0% (0.0%, 29.9%)  50.0% (15.0%, 85.0%)  25.0% (4.6%, 69.9%)  81.8% (52.3%, 94.9%)  0.0% (0.0%, 49.0%) | 0.0% (0.0%, 29.9%)  75.0% (30.1%, 95.4%)  50.0% (15.0%, 85.0%)  90.0% (62.3%, 98.4%)  0.0% (0.0%, 49.0%) |
| Stomach | Stage I (5)  Stage II (7)  Stage III (7)  Stage IV (9)  Unknown (2) | 20.0% (3.6%, 62.4%)  57.1% (25.0%, 84.2%)  85.7% (48.7%, 97.4%)  100.0% (70.1%, 100.0%)  50.0% (9.5%, 90.5%) | 40.0% (11.8%, 76.9%)  57.1% (25.0%, 84.2%)  85.7% (48.7%, 97.4%)  100.0% (70.1%, 100.0%)  50.0% (9.5%, 90.5%) |
| Bladder and Urinary | Stage I (7)  Stage II (9)  Stage III (4)  Stage IV (6)  Unknown (2) | 14.3% (2.6%, 51.3%)  44.4% (18.9%, 73.3%)  75.0% (30.1%, 95.4%)  66.7% (30%, 90.3%)  50.0% (9.5%, 90.5%) | 28.6% (8.2%, 64.1%)  66.7% (35.4%, 87.9%)  100.0% (51.0%, 100.0%)  100.0% (61.0%, 100.0%)  50.0% (9.5%, 90.5%) |
| Esophagus | Stage I (6)  Stage II (5)  Stage III (8)  Stage IV (7)  Unknown (1) | 0.0% (0.0%, 39.0%)  20.0% (3.6%, 62.4%)  75.0% (40.9%, 92.9%)  85.7% (48.7%, 97.4%)  100.0% (20.7%, 100.0%) | 33.3% (9.7%, 70.0%)  40.0% (11.8%, 76.9%)  75.0% (40.9%, 92.9%)  85.7% (48.7%, 97.4%)  100.0% (20.7%, 100.0%) |
| Liver and Bile Duct | Stage I (5)  Stage II (6)  Stage III (8)  Stage IV (5)  Unknown (1) | 40.0% (11.8%, 76.9%)  83.3% (43.6%, 97%)  100.0% (67.6%, 100.0%)  100.0% (56.6%, 100.0%)  0.0% (0.0%, 79.3%) | 40.0% (11.8%, 76.9%)  83.3% (43.6%, 97.0%)  100.0% (67.6%, 100.0%)  100.0% (56.6%, 100.0%)  0.0% (0.0%, 79.3%) |
| Anus | Stage I (1)  Stage II (8)  Stage III (3)  Stage IV (3)  Unknown (1) | 0.0% (0.0%, 79.4%)  62.5% (30.6%, 86.3%)  66.7% (20.8%, 93.9%)  100.0% (43.9%, 100.0%)  0.0% (0.0%, 79.3%) | 0.0% (0.0%, 79.4%)  75.0% (40.9%, 92.9%)  66.7% (20.8%, 93.9%)  100.0% (43.9%, 100.0%)  0.0% (0.0%, 79.3%) |
| Ovary | Stage I (1)  Stage II (1)  Stage III (7)  Stage IV (3)  Unknown (2) | 0.0% (0.0%, 79.4%)  0.0% (0.0%, 79.4%)  85.7% (48.7%, 97.4%)  66.7% (20.8%, 93.9%)  100.0% (34.2%, 100.0%) | 0.0% (0.0%, 79.4%)  0.0% (0.0%,79.4%)  85.7% (48.7%, 97.4%)  66.7% (20.8%, 93.9%)  100.0% (34.2%, 100.0%) |
| Vulva | Stage I (6)  Stage II (0)  Stage III (3)  Stage IV (20  Unknown (2) | 0.0% (0.0%, 39.0%)  -  66.7% (20.8%, 93.9%)  100.0% (34.2%, 100.0%)  50.0% (9.5%, 90.5%) | 16.7% (3.0%, 56.4%)  -  66.7% (20.8%, 93.9%)  100.0% (34.2%, 100.0%)  50.0% (9.5%, 90.5%) |
| Thyroid | Stage I (4)  Stage II (4)  Stage III (0)  Stage IV (4)  Unknown (1) | 0.0% (0.0%, 49.0%)  0.0% (0.0%, 49.0%)  -  50.0% (15.0%, 85.0%)  0.0% (0.0%, 79.3%) | 0.0% (0.0%, 49.0%)  0.0% (0.0%, 49.0%)  -  75.0% (30.1%, 95.4%)  0.0% (0.0%, 79.3%) |
| Cervix uteri | Stage I (2)  Stage II (3)  Stage III (3)  Stage IV (2)  Unknown (3) | 100.0% (34.2%, 100%)  66.7% (20.8%, 93.9%)  100.0% (43.9%, 100.0%)  100.0% (34.2%, 100.0%)  33.3% (6.1%, 79.2%) | 100.0% (34.2%, 100%)  66.7% (20.8%, 93.9%)  100.0% (43.9%, 100.0%)  100.0% (34.2%, 100.0%)  33.3% (6.1%, 79.2%) |
| Small Intestine | Stage I (0)  Stage II (3)  Stage III (4)  Stage IV (3)  Unknown (0) | -  66.7% (20.8%, 93.9%)  25.0% (4.6%, 69.9%)  33.3% (6.1%, 79.2%)  - | -  66.7% (20.8%, 93.9%)  25.0% (4.6%, 69.9%)  33.3% (6.1%, 79.2%)  - |
| NHL | Stage I (1)  Stage II (0)  Stage III (2)  Stage IV (4)  Unknown (0) | 0.0% (0.0%, 79.3%)  -  0.0% (0.0%, 65.8%)  50.0% (15.0%, 85.0%)  - | 0.0% (0.0%, 79.3%)  -  50.0% (9.5%, 90.5%)  75.0% (30.1%, 95.4%)  - |
| Testis | Stage I (2)  Stage II (0)  Stage III (0)  Stage IV (0)  Unknown (0) | 50.0% (9.5%, 90.5%)  -  -  -  - | 50.0% (9.5%, 90.5%)  -  -  -  - |
| Multiple Myeloma | Stage I (0)  Stage II (1)  Stage III (0)  Stage IV (0)  Unknown (0) | -  0.0% (0.0%, 79.3%)  -  -  - | -  30.0% (0.0%, 79.%)  -  -  - |

MP, Methylation-Protein; NHL, Non-Hodgkin Lymphoma. *MP V1 Specificity=98.5% (95% CI: 97.9-98.9%) **MP V2 Specificity=97.4% (95% CI: 96.7-97.9%).

Table S3. MP V2 clinical validation set organ type and stage distribution

|  | **I**  **N n** | | **II**  **n** | **III**  **n** | **IV**  **n** |
| --- | --- | --- | --- | --- | --- |
| Prostate | 51 | 12 | 21 | 11 | 7 |
| Breast | 50 | 38 | 6 | 4 | 2 |
| Lung and Bronchus | 57 | 11 | 11 | 14 | 21 |
| Colon and Rectal | 38 | 7 | 10 | 13 | 8 |
| Kidney | 19 | 8 | 3 | 5 | 3 |
| Uterus | 17 | 9 | 2 | 4 | 2 |
| Head and Neck | 17 | 4 | 6 | 3 | 4 |
| Pancreas | 17 | 2 | 5 | 3 | 7 |
| Bladder and Urinary | 12 | 3 | 6 | 2 | 1 |
| Liver and Bile Duct | 12 | 3 | 4 | 3 | 2 |
| Thyroid | 5 | 4 | 1 | 0 | 0 |
| Stomach | 7 | 2 | 2 | 1 | 2 |
| Ovary | 7 | 1 | 1 | 3 | 2 |
| Esophagus | 6 | 1 | 2 | 1 | 2 |
| Small Intestine | 3 | 0 | 1 | 0 | 2 |
| Cervix Uteri | 2 | 0 | 1 | 1 | 0 |
| Anus | 2 | 0 | 1 | 1 | 0 |
| Vulva | 1 | 1 | 0 | 0 | 0 |
| Testis | 1 | 0 | 0 | 1 | 0 |
| Total (excluding breast, prostate) | 223 | 56 | 56 | 55 | 56 |
| Total (all types) | 324 | 106 | 83 | 70 | 65 |

MP, Methylation-Protein

Table S4. MP V2 classifier sensitivity by organ type and stage in the clinical validation set

| **Organ Type** | **Sensitivity**  **(95% CI)**  **n/N** | | | | |
| --- | --- | --- | --- | --- | --- |
|  | **Overall** | **Stage I** | **Stage II** | **Stage III** | **Stage IV** |
| Prostate (n=51) | 3.9%  (1.1-13.2%)  2/51 | 0.0%   (0.0-24.2%)  0/12 | 0.0%   (0.0-15.5%)  0/21 | 0.0%   (0.0-25.9%)  0/11 | 28.6%   (8.2-64.1%)  2/7 |
| Breast  (n=50) | 16.0%  (8.3-28.5%)  8/50 | 5.3%   (1.5-17.3%)  2/38 | 33.3%   (9.7-70%)  2/6 | 50%   (15.0- 85.0%)  2/4 | 100%   (34.2-100.0%)  2/2 |
| Lung & Bronchus  (n=57) | 59.6%  (46.7-71.4%)  34/57 | 27.3%   (9.7-56.6%)  3/11 | 45.5%   (21.3-72%)  5/11 | 64.3%   (38.8-83.7%)  9/14 | 81%   (60.0-92.3%)  17/21 |
| Colon & Rectal  (n=38) | 60.5%  (44.7-74.4%)  23/38 | 14.3%   (2.6-51.3%)  1/7 | 60%  (31.3-83.2%)  6/10 | 61.5%  (35.5-82.3%)  8/13 | 100%   (67.6-100.0%)  8/8 |
| Kidney  (n=19) | 26.3%  (11.8-48.8%)  5/19 | 0.0%   (0.0-32.4%)  0/8 | 0.0%   (0.0-56.1%)  0/3 | 40.0%   (11.8-76.9%)  2/5 | 100.0%   (43.9-100.0%)  3/3 |
| Head & Neck (n=17) | 58.8%  (36.0-78.4%)  10/17 | 50.0%   (15.0- 85.0%)  2/4 | 33.3%   (9.7-70.0%)  2/6 | 100.%   (43.9-100.0%)  3/3 | 75.0%  (30.1-95.4%)  3/4 |
| Pancreas  (n=17) | 70.6%  (46.9-86.7%)  12/17 | 100.0%   (34.2-100.0%)  2/2 | 20.0%   (3.6-62.4%)  1/5 | 66.7%   (20.8-93.9%)  2/3 | 100.0%   (64.6-100.0%)  7/7 |
| Uterus  (n=17) | 17.6%  (6.2-41.0%)  3/17 | 11.1%  (2.0-43.5%)  1/9 | 50.0%   (9.5-90.5%)  1/2 | 0.0%  (0.0-49.0%)  0/4 | 50.0%  (9.5-90.5%)  1/2 |
| Liver & Bile Duct (n=12) | 91.7%  (64.6-98.5%)  11/12 | 66.7%  (20.8-93.9%)  2/3 | 100.0%  (51-100.0%)  4/4 | 100.0%   (43.9-100.0%)  3/3 | 100.0%   (34.2-100.0%)  2/2 |
| Bladder & Urinary (n=12) | 16.7%  (4.7-44.8%)  2/12 | 33.3%   (6.1-79.2%)  1/3 | 0.0%   (0.0-39.0%)  0/6 | 0.0%   (0.0-65.8%)  0/2 | 100.0%   (20.7-100.0%)  1/1 |
| Ovary  (n=7) | 100.0%  (64.6-100.0%)  7/7 | 100.0%  (20.7-100.0%)  1/1 | 100.0%   (20.7-100.0%)  1/1 | 100.0%   (43.9-100.0%)  3/3 | 100.0%   (34.2-100.0%)  2/2 |
| Stomach  (n=7) | 57.1%  (25.0-84.2%)  4/7 | 0.0%   (0.0- 65.8%)  0/2 | 50.0%   (9.5-90.5%)  1/2 | 100.0%   (20.7-100.0%)  1/1 | 100.0%   (34.2-100.0%)  2/2 |
| Esophagus  (n=6) | 66.7%  (30.0-90.3%)  4/7 | 0.0%   (0.0-79.3%)  0/1 | 50.0%   (9.5-90.5%)  1/2 | 100.0%   (20.7-100.0%)  1/1 | 100.0%   (34.2-100.0%)  2/2 |
| Thyroid  (n=5) | 20.0%  (3.6-62.4%)  1/5 | 25.0%   (4.6-69.9%)  1/4 | 0.0%   (0.0-79.3%)  0/1 |  |  |
| Small Intestine (n=3) | 100.0%  (43.9-100.0%)  3/3 |  | 100.0%  (20.7-100.0%)  1/1 |  | 100.0%   (34.2-100.0%)  2/2 |
| Anus  (n=2) | 100.0%  (34.2-100.0%)  2/2 |  | 100.0%   (20.7-100.0%)  1/1 | 100.0%   (20.7-100.0%)  1/1 |  |
| Cervix Uteri  (n=2) | 50.0%  (9.5- 90.5%)  1/2 |  | 0.0%   (0.0-79.3%)  0/1 | 100.0%   (20.7-100.0%)  1/1 |  |
| Vulva  (n=1) | 100.0%  (20.7-100.0%)  1/1 | 100.0%   (20.7-100.0%)  1/1 |  |  |  |
| Testis  (n=1) | 100.0%  (20.7-100.0%)  1/1 |  |  | 100.0%   (20.7-100.0%)  1/1 |  |

MP, Methylation-Protein.

Table S5. Summary of MP V2 detected tumor types and histological subtypes

| **Tumor Type** | **Histology Subtype** |
| --- | --- |
| Anus | Squamous Cell Carcinoma |
|  | Adenocarcinoma |
| Bladder and Urinary | Invasive Urothelial Carcinoma |
|  | Squamous Cell Carcinoma |
| Breast | Invasive Ductal Carcinoma |
|  | Invasive Lobular Carcinoma |
|  | Tubular Carcinoma |
|  | Mucinous Adenocarcinoma |
|  | Breast Carcinoma with Apocrine Features |
| Cervix | Squamous Cell Carcinoma |
|  | Adenocarcinoma |
| Colon and Rectum | Rectal adenocarcinoma |
|  | Colon adenocarcinoma |
|  | Neuroendocrine Tumor  Squamous Cell carcinoma |
| Esophagus | Adenocarcinoma |
|  | Neuroendocrine carcinoma |
|  | Squamous Cell Carcinoma |
| Head and Neck | Oropharynx (p16+) |
|  | Oropharynx (p16-) |
|  | Nasopharynx |
|  | Hypopharynx |
|  | Oral Cavity |
|  | Larynx |
| Kidney | Clear-Cell Renal Cell Carcinoma |
|  | Chromophobe Renal Cell Carcinoma |
| Renal Pelvis | Sarcomatoid Carcinoma |
|  | Adenocarcinoma |
|  | Urothelial Carcinoma |
| Liver and Bile Duct | Hepatocellular Carcinoma |
|  | Cholangiocarcinoma |
| Lung and Bronchus | Adenocarcinoma |
|  | Large Cell Carcinoma |
|  | Squamous Cell Carcinoma |
|  | Small Cell Carcinoma |
|  | Carcinoid Tumor |
| Non-Hodgkin Lymphoma | B-Cell Lymphoma |
| Ovary | Serous Carcinoma |
|  | Mucinous Adenocarcinoma |
| Pancreas | Non- Pancreatic Ductal Adenocarcinoma Exocrine Pancreatic Tumors |
|  | Pancreatic Ductal Adenocarcinoma |
|  | Neuroendocrine Tumors (NETs) |
| Prostate | Adenocarcinoma |
| Small Intestine | Adenocarcinoma |
|  | Neuroendocrine Carcinoma |
| Stomach | Adenocarcinoma |
|  | Neuroendocrine Carcinoma |
| Testis | Seminoma |
| Thyroid | Anaplastic Thyroid Carcinoma |
|  | Papillary Thyroid Carcinoma |
| Uterus | Endometrioid Carcinoma |
|  | Carcinosarcoma |
|  | Serous Carcinoma |
|  | Squamous Cell Carcinoma |
|  | Clear Cell Carcinoma |
| Vulva | Squamous Cell Carcinoma |

MP, Methylation-Protein.

Figure S1. ASCEND-2 test set overall sensitivity by organ type for MP V1* (gray) and MP V2** (blue) classifiers

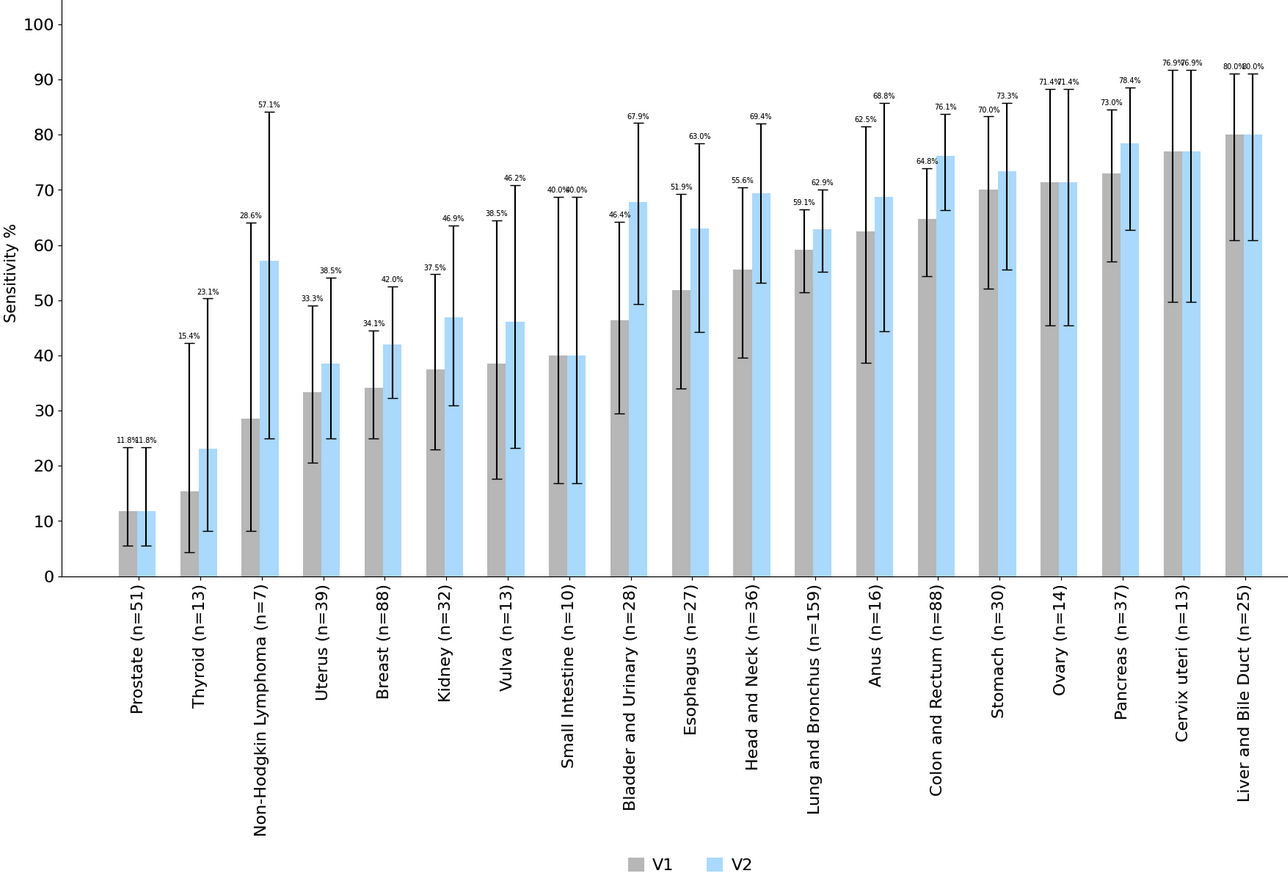

*MP V1 Specificity=98.5% (95% CI: 97.9-98.9%) **MP V2 Specificity=97.4% (95% CI: 96.7-97.9%).
